# Mass testing for discovery and control of COVID-19 outbreaks in adult social care: an observational study and cost-effectiveness analysis of 14,805 care homes in England

**DOI:** 10.1101/2024.06.06.24308563

**Authors:** Siyu Chen, Richard Creswell, Rachel Hounsell, Liberty Cantrell, Sumali Bajaj, Prabin Dahal, Lok Hei Tsui, Olumide Kolade, Ma’ayan Amswych, Reshania Naidoo, Tom Fowler, Susan Hopkins, Kasia Stepniewska, Merryn Voysey, Lisa White, Rima Shretta, Ben Lambert, the EY-Oxford Health Analytics Consortium

## Abstract

**Introduction:** We retrospectively evaluated the impact of COVID-19 testing among residents and staff in social care homes in England.

**Methods:** We obtained 80 million reported polymerase chain reaction (PCR) and lateral flow device (LFD) test results, from 14,805 care homes (residents and staff) in England, conducted between October 2020 and March 2022. These testing data were then linked to care home characteristics, test costs and 24,500 COVID-19-related deaths of residents. We decomposed the mechanism of outbreak mitigation into outbreak discovery and outbreak control and used Poisson regressions to investigate how reported testing intensity was associated with the size of outbreak discovered and to uncover its association with outbreak control. We used negative binomial regressions to determine the factors influencing COVID- 19-related deaths subsequent to outbreaks. We performed a cost-effectiveness analysis of the impact of testing for preventing COVID-19-related deaths of residents.

**Results:** Reported testing intensity generally reflected changes in testing policy over time, although there was considerable heterogeneity among care homes. Client type was the strongest determinant of whether COVID-19-related deaths in residents occurred subsequent to testing positive. Higher staff-to-resident ratios were associated with larger outbreak sizes but rapid outbreak control and a decreased risk of COVID-19-related deaths. Assuming our regression estimates represent causal effects, care home testing in England was cost effective at preventing COVID-19-related deaths among residents during the pandemic and approximately 3·5-times more cost effective prior to the vaccine rollout.

**Conclusions:** PCR and LFD testing was likely an impactful intervention for detecting and controlling COVID-19 outbreaks in care homes in England and cost effective for preventing COVID-19- related deaths among residents. In future pandemics, testing must be prioritised for care homes, especially if severe illness and death particularly affects older people or individuals with characteristics similar to care home residents, and an efficacious vaccine is unavailable.

**Key Messages:** *Evidence before this study:* Mass diagnostic testing plays a key role in any pandemic response. COVID-19 testing in the adult social care sector in England was implemented by NHS Test and Trace (later the UK Health Security Agency, UKHSA). Prior to the large-scale evaluation we report here, a pilot testing evaluation programme had been conducted in Liverpool.

*Added value of this study:* This study represents the first large-scale evaluation of England’s COVID-19 testing programme in adult social care homes. It encompasses data from residents and staff from 14,805 adult social care homes in England. Our findings show that testing was an important and useful public health intervention that had a considerable impact. It was also cost effective at preventing COVID-19-related deaths in adult social care home residents.

*Implications of all the available evidence:* Our study has implications for the development of testing policies in adult social care homes in any future pandemic, particularly if it involves a respiratory disease similar to COVID-19. We found that while testing was a crucial public health intervention in adult social care homes, there were also large heterogeneities seen among care homes. Policymakers thus need to consider whether a one-size-fits-all policy for care home testing is the most effective approach to take in the face of any future pandemic.

## Introduction

Adult social care homes were hit hard by the time-varying epidemic waves of COVID-19, both in the UK and internationally ^1–5^, experiencing high levels of COVID-19-related mortality (and excess mortality) among residents and staff.^6–12^ This was in part associated with old age, frailty, and comorbidities of care home residents, who typically have multiple needs and require a range of frequent close care and support interactions.^13–15^ It was also partly associated with the close contact among residents and between residents and staff, with staff usually living in the wider community, frequently working in more than one care home, and potentially carrying infections as they moved between care homes, resulting in rapid transmission and subsequent severe outbreaks.^16^ A consensus statement on the association between the discharge of patients from hospitals and COVID-19 in care homes reported that ‘at least some care home outbreaks were caused or partly caused or intensified by discharges from hospital in the UK’^17^.

In response to the COVID-19 pandemic, the UK government committed to mass testing, with initial testing commencing in May 2020 ^18^. NHS Test and Trace (NHSTT) was a service within the Department of Health and Social Care (DHSC); it was moved into the UK Health Security Agency which is an executive agency. One of the first established testing services was within the adult social care sector.^19^ England’s COVID-19 testing policy within adult social care (which encompasses care homes) had several intended objectives, including to control the spread of infection^20^ and consequently reduce hospitalisation and mortality rates. However, the testing policy and its implementation changed multiple times throughout the course of the COVID-19 pandemic and depending on the availability of tests. For staff testing, routine weekly asymptomatic polymerase chain reaction (PCR) testing was rolled out in October 2020 and discontinued from February 2022.^21^ This was enhanced by twice-weekly testing with lateral flow devices (LFDs) in December 2020,^22^ ^23^ increasing to three times a week in December 2021 in response to the emergence of the Omicron variant,^24^ and further increasing to daily pre-shift LFDs in February 2022.^25^ For resident testing, a policy of monthly PCR testing was maintained until February 2022,^21^ with increased frequency during outbreaks in care homes (involving both LFD and PCR tests, including for staff) from July 2020 (Supplementary Materials Section 1 and Supplementary Table S1).^26–28^ However, the impacts of this large-scale COVID-19 testing programme and associated policies in social care settings have yet to be evaluated.

Studies of the impact of the COVID-19 pandemic on care homes have tended to focus on describing correlations between the characteristics of a care home with infection rates,^29^ outbreak magnitudes,^6^ ^12^ ^30^ ^31^ or COVID-19-related deaths.^6^ ^32^ These studies were generally conducted early on during the pandemic, when COVID-19 tests were extremely scarce, or they included very few care homes. Individual-level risk factors for care home residents testing positive for SARS-CoV-2 in Wales (UK)^15^ and infection severity in the U.S.^33–36^ were also examined. Here, we describe an in-depth analysis of the impact of testing programmes for COVID-19 in adult social care homes, which formed part of a retrospective, independent evaluation of the COVID-19 mass testing programme between October 2020 and March 2022 in England, commissioned by the UK Health Security Agency (UKHSA).^19^ ^37^

## Materials and Methods

### Data sources

As part of a retrospective evaluation of England’s COVID-19 testing policy^19^, we had access to mass testing data from care homes, which we later linked to testing cost, mortality, and care home characteristics data. This enabled us to assess the factors associated with the level of testing reported in care homes and probe the mechanisms by which testing affected outbreak discovery and control and COVID-19-related deaths in care homes. Furthermore, we compared the cost effectiveness of counterfactual SARS-CoV-2 testing intensity levels in care homes on resident quality-adjusted life-years (QALYs) gained. We were supplied “Pillar 2” testing data for both PCR and LFD tests (Pillar 2 refers to swab testing for SARS-CoV-2 in the general population, through commercial partnerships, either processed in a laboratory or more rapidly via LFDs.^38^) Our study involved 14,805 individual care homes across our study period. According to carehome.co.uk, there are currently 16,726 care homes in the UK, of which 14,228 are in England^39^. Our analysis period covered 1 October 2020 to 30 March 2022. The testing data were linked to several other datasets, either at the individual care home level or at the level of the lower-tier local authority (LTLA) encompassing the care homes, including:

- COVID-19-related deaths of care home residents as reported to the Care Quality Commission (CQC), which included both confirmed and suspected COVID-19-related deaths.
- Characteristics of individual care homes obtained from the NHS North of England Care System Support (NECS) Capacity Tracker,^40^ including the CQC rating and primary client type served by the care home (Table 1).
- Weekly estimates of COVID-19 community prevalence according to the LTLA within which each care home was situated. These estimates were generated using a causal debiasing methodology,^41^ which used high-quality randomised surveillance data of swab-positive results provided via the REACT-1 survey^42^ to debias PCR testing data obtained through Pillar 2^2^.
- Weekly vaccination data for staff and residents in individual care homes by dose type (dose 1, dose 2, dose 3, booster, spring 2022 booster, or null).
- Weekly proportions of SARS-CoV-2 lineages by LTLA, downloaded from the Sanger Institute website.^43^
- Mid-2020 estimates of population sizes by region in England, obtained from the publicly accessible Office for National Statistics (ONS) website.^44^
- COVID-19 testing costs derived from the Cost Allocation Project (an unpublished internal project) conducted by UKHSA for the ONS. These testing cost data were primarily sourced from UKHSA general ledgers.

**Table 1.** Characteristics of care homes studied.

|  |  | Care home size including staff and residents<sup>*</sup> | Number of staff members per resident (staff-to-resident ratio)<sup>*</sup> | Number of reported PCR + LFD tests per residents per week<sup>*</sup> | Number of reported PCR + LFD tests per staff per week<sup>*</sup> | Number (%) of care homes | Total number (%) of COVID-19 deaths reported (suspected + confirmed) |
| --- | --- | --- | --- | --- | --- | --- | --- |
| CQC rating | Inadequate | 37.0<br>(24.0, 59.0) | 1.0<br>(0.6, 1.2) | 2.5<br>(1.7, 3.5) | 12.2<br>(7.1, 19.3) | 302 (2.0%) | 569 (2.3%) |
|  | Requires improvement | 45.0<br>(28.0, 68.0) | 1.0<br>(0.7, 1.3) | 3.1<br>(2.3, 4.1) | 18.1<br>(11.6, 26.4) | 2,421 (16.4%) | 5,065 (20.7%) |
|  | Good | 39.0<br>(22.0, 63.0) | 0.9<br>(0.5, 1.3) | 3.3<br>(2.4, 4.4) | 19.8<br>(12.7, 28.3) | 11,197 (75.6%) | 17,619 (71.9%) |
|  | Outstanding | 52.0<br>(29.0, 86.0) | 0.8<br>(0.4, 1.1) | 4.0<br>(2.9, 5.3) | 23.4<br>(17.0, 34.1) | 634 (4.3%) | 1,093 (4.5%) |
|  | Null or unrated | 40.0<br>(23.0, 65.0) | 0.9<br>(0.5, 1.2) | 3.3<br>(2.4, 4.4) | 19.4<br>(12.4, 27.9) | 251 (1.7) | 154 (0.6%) |
|  | Total |  |  |  |  | 14,805 | 24,500 |
| Primary client | Older people (≥65 years) | 52.0 | 1.1<br>(0.9, 1.3) | 3.4<br>(2.7, 4.3) | 22.8<br>(15.2, 32.0) | 7,020 (47.4%) | 17,839 (72.8%) |
|  | Learning disability | 20.0<br>(14.0, 30.0) | 0.4<br>(0.3, 0.6) | 3.0<br>(1.6, 4.7) | 15.3<br>(9.5, 22.0) | 4,233 (28.6%) | 285 (1.2%) |
|  | Dementia | 64.0<br>(42.0, 91.0) | 1.1<br>(0.9, 1.3) | 3.3<br>(2.6, 4.2) | 21.0<br>(14.3, 29.4) | 1,881 (12.7%) | 5,699 (23.3%) |
|  | Mental health | 21.0<br>(14.0, 31.0) | 0.9<br>(0.6, 1.3) | 3.2<br>(2.2, 4.8) | 15.8<br>(8.8, 23.6) | 917 (6.2%) | 192 (0.8%) |
|  | Physical disability | 47.0<br>(33.0, 66.0) | 0.6<br>(0.5, 1.0) | 3.6<br>(2.7, 4.6) | 19.5<br>(13.8, 30.2) | 269 (1.8%) | 198 (0.8%) |
|  | Other | 40.0<br>(23.0, 65.0) | 0.9<br>(0.5, 1.2) | 3.3<br>(2.4, 4.4) | 19.5<br>(12.4, 28.1) | 485 (3.3%) | 287 (1.2%) |
|  | Total |  |  |  |  | 14,805 | 24,500 |
| Location type | Social care organisation | 40.0<br>(23.0, 65.0) | 0.9<br>(0.5, 1.2) | 3.3<br>(2.4, 4.4) | 19.6<br>(12.5, 28.1) | 14,604 (98.6%) | 24,453 (99.8%) |
|  | Independent | 83.0<br>(35.0, 90.0) | 0.8<br>(0.5, 1.0) | 2.6<br>(1.4, 4.7) | 25.5<br>(8.8, 39.0) | 10 (0.07%) | 21 (0.09%) |
|  | NHS healthcare | 20.0<br>(18.0, 43.0) | 0.5 | 3.6<br>(2.8, 6.4) | 10.0<br>(6.3, 17.3) | 18 (0.12%) | 12 (0.05%) |
|  | Other | 40.0<br>(23.0, 65.0) | 0.9<br>(0.5, 1.2) | 3.3<br>(2.4, 4.4) | 19.5<br>(12.5, 28.1) | 173 (1.2%) | 14 (0.06%) |
|  | Total |  |  |  |  | 14,805 | 24,500 |
| Care home characteristics<sup>note</sup> | Nursing home | 68.0<br>(48.0, 94.0) | 1.1<br>(0.9, 1.4) | 3.4<br>(2.8, 4.3) | 24.7<br>(16.1, 35.0) | 4,176 (28.2%) | 13,222 (54.0%) |
|  | Residential home | 32.0<br>(19.0, 49.0) | 0.8<br>(0.4, 1.1) | 3.2<br>(2.3, 4.4) | 17.9<br>(11.4, 25.5) | 10,458 (70.6%) | 11,264 (46.0%) |
|  | Community home | 40.0<br>(22.0, 68.0) | 0.6<br>(0.4, 1.0) | 3.5<br>(2.4, 5.0) | 18.9<br>(12.0, 26.4) | 523 (3.5%) | 660 (2.7%) |

**Note:** According to the Care Quality Commission (CQC), community healthcare services (CHC) supply a range of healthcare staff other the doctors, for example, nurses or allied health professionals, to people who need healthcare support in their own home, in community settings or in child development units<sup>48</sup>. 'Acute care home' provide medical and/or surgical investigations, diagnosis and treatment for physical illness or condition, injury or disease. According to carehome.co.uk, 'residential care homes' offer emergency, respite, short term, long-term care and even palliative care to older people and young adults who stay in a residential setting rather than in their own home or family home. 'Nursing care homes' offer the same type of care as residential ones but with the addition of 24-hour medical care from a qualified nurse<sup>49</sup>.

More details on data source and cleaning can be found in the Supplementary Materials Section 2-3 and Supplementary Table S1.

### Patient and public involvement

As part of the COVID-19 testing initiative evaluation programme^45^, addressing the issue of inequalities in COVID-19 testing engaged heavily with patient and public. For example, evaluations of LFD and PCR test user experience were conducted among blind and partially sighted people by the Home Test Service in collaboration with relevant stakeholders^46^; assessment of general public testing behaviours was implemented through fortnightly opinion surveys^47^. We presented care home providers basic statistic findings to help to encourage familiarity with research concepts and terminology. Stakeholders from care home organisations highlighted the ‘very short notice that was received of the move to twice-weekly LFD testing, in December 2020, as being particularly challenging to implement in a short space of time, during what was also the Christmas period.’ (more details see page 142 of reference ^19^) Stakeholders from UKHSA and DHSC described the barriers, for example, ‘in general, the workforce in care homes was not trained to take swabs, and this initially presented a considerable challenge ’ (more details see page 142 of reference^19^) and motivations of implementing the testing policy, reporting the testing results, and acting on a positive result in practice during the COVID-19 pandemic. We expanded the evaluation scope and refined the research findings to align them better with things that care providers cared about the most. Through these iterative processes, care home providers made high value contributions to both the design and the interpretation of the study, while we retained the rigors of the scientific work.

## Methods

We define reported testing intensity over a particular time period, e.g. weekly, of a particular group, e.g. staff or resident counts, in a particular care home as:

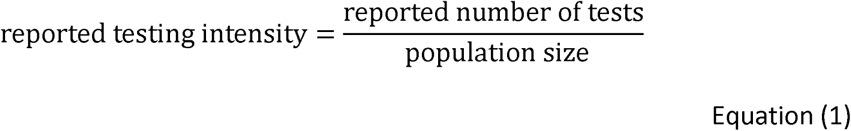

We assumed that the sole impact of testing on resident deaths in care homes was through its impact on COVID-19 outbreak discovery and outbreak control. By testing staff and residents more frequently at higher intensities, outbreaks (or single, isolated cases) can be discovered earlier, with the result that initial outbreak sizes upon detection are smaller (Fig. 1(A)). Once an outbreak has been discovered, higher testing coverages enable cases to be identified earlier and isolated more effectively, resulting in more rapid reductions in onward cases and thus more rapid control (Fig. 1(B)). Through both mechanisms, higher testing coverages would lead to smaller outbreaks of COVID-19 in care homes and likely a reduction in the number of deaths among residents.

**Fig. 1.**
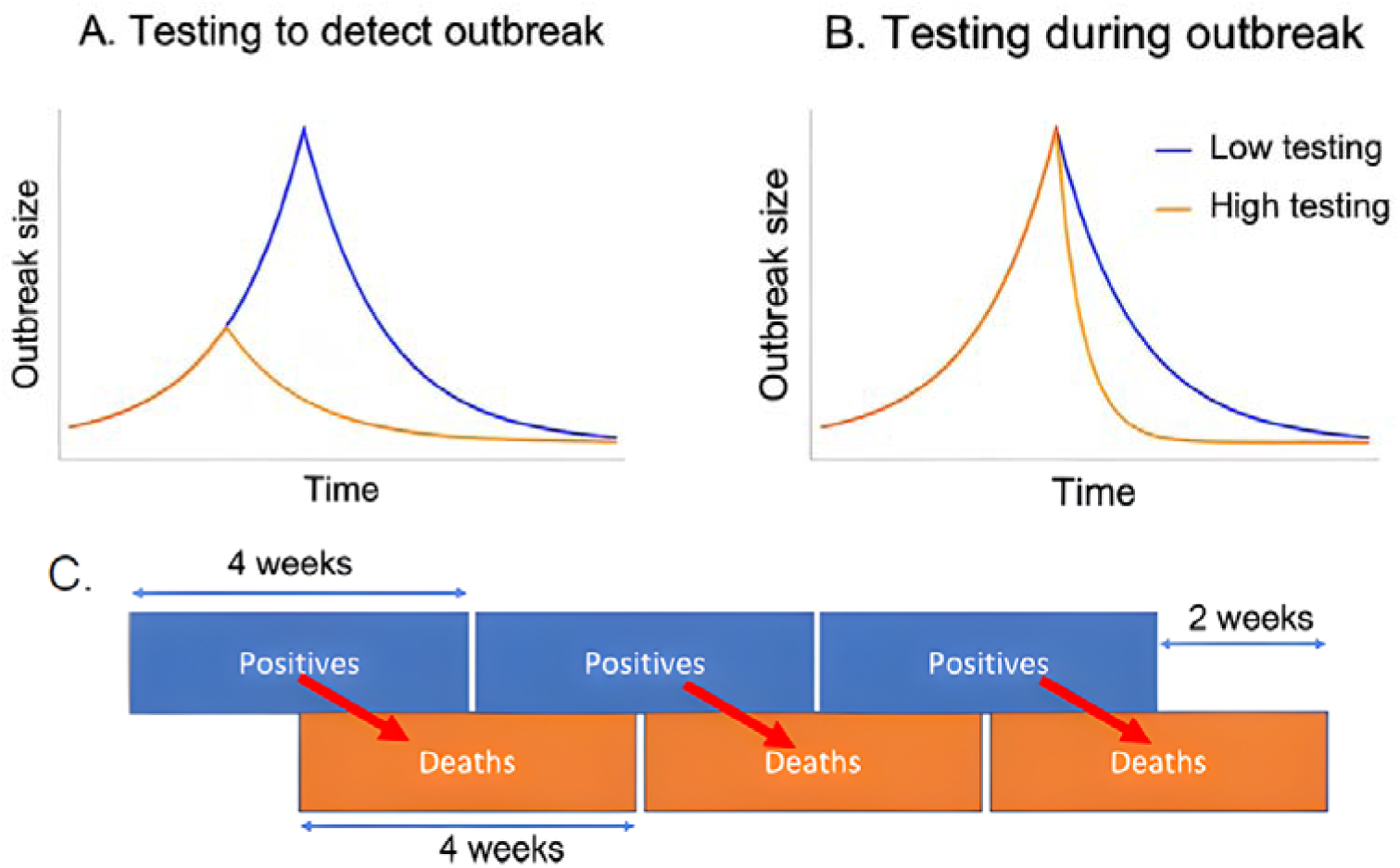
Characterising the two pathways through which our models assume testing influences outbreak dynamics (A and B) and design of positives-to-deaths regression (C). Panel (A) shows that higher levels of testing are associated with outbreaks being detected earlier, when they are smaller in size (leading to more rapid outbreak control). Panel (B) shows that higher levels of testing during outbreaks are associated with more rapid control of outbreaks. Panel (C) shows the link between our dependent variable (COVID-19-related deaths) and testing variables, one of our independent variables, in our regressions that focused on COVID-19 deaths in individuals who had received a positive test result for COVID-19.

To test this hypothesis, we used Poisson regression models to empirically probe the mechanisms by which changes in testing influenced the number of positive cases. We also used negative binomial regressions, to probe the relationship between positive cases and COVID-19-related deaths.

### The association between reported testing intensity and the size of outbreaks when discovered

We used Poisson regressions to investigate how testing intensity in both residents and staff influenced outbreak size during the process of “outbreak discovery”. We define outbreak discovery as the period when care homes uncovered no positive cases one week but did so in the following week. These regressions took the general form:

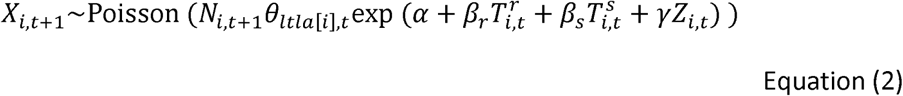

where *X_i,t_*, denotes the number of positive tests in care home *i* and week *t*, and *N_i,t_*, denotes the corresponding number of tests conducted; *θ_itla_*_[*a*],*t*_, denotes the estimated COVID-19 prevalence in the LTLA that encompasses care home *i*; *T^r^_i,t_* and *T^s^_i,t_* denote the reported testing intensities in residents and staff, respectively; and *Z_i,t_*, denotes a vector of additional covariates, including weekly time dummies that account for secular changes in the relationship between tests and positive results over time across England. In what follows, we group PCR and LFD tests together for either residents or staff, as these regression results tended to be more stable. More details can be found in Supplementary Materials Section 4-5.

### The association between reported testing intensity and outbreak control

We also used Poisson regressions to investigate how, after positive cases were found within a care home, the response was able to identify (and, if possible, isolate) cases, leading to reductions in the size of the outbreaks in subsequent weeks. We refer to this process as “outbreak control”. To do so, we considered only those weeks where the previous week had at least one positive case in either staff or residents. The regressions using these data then took the following form:

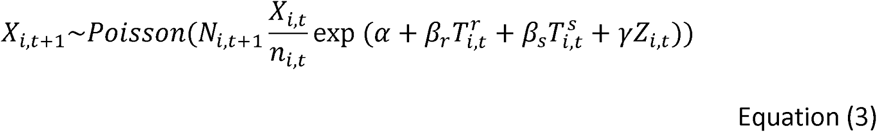

where the variables appearing both here and in Equation (2) have common meanings, while *n_it_* > 0 is the total number of members of care home *i* at time *t* (i.e. the sum of residents and staff numbers). More details can be found in Supplementary Materials Section 4 and 6.

### COVID-19-related deaths of residents

We investigated the factors influencing whether positive COVID-19 cases in residents became deaths by considering four-week blocks for aggregating positive results and deaths (which were chosen to overlap by two weeks in the middle, with future deaths depending on the previous cases), to account for the typical delay between a case being detected and death. This is illustrated in Fig. 1(C).

The regressions took the form:

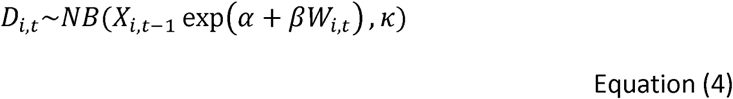

where NB(*μ*, *κ*) denotes a negative binomial distribution parameterised to have mean *µ* > 0 and an overdispersion parameter *κ* where, as *κ* → ∞, the distribution tends to a Poisson; *D_it_* > 0 denotes the count of COVID 19-related deaths in a given block, *t*; *X_it_*_-1_ > 0 denotes the positive count in residents only in the previous block; and *W_i,t_*, represents various covariates that may affect whether positive test results predict deaths. More details can be found in Supplementary Materials Section 7.

### Deaths under hypothetical testing scenarios

We used the fitted regression models describing outbreak discovery (Equation (2)) and outbreak control (Equation (3)), together with the model for total COVID-19-related deaths among residents (Equation (4)), to estimate the number of such deaths that would occur under counterfactual testing rates of 50%, 75%, 125%, 150%, and 200% of the actual testing volume. This enabled us to assess the impact of increased or decreased rates of testing and how close the testing regimen actually implemented was to a theoretical “optimal” testing regimen. In using our regression results in this way, we may tentatively assume that our estimates represent causal effects. More details can be found in Supplementary Materials Section 8-10.

### Cost-effectiveness analysis

We conducted a cost-effectiveness analysis to evaluate the impact of testing in care homes on mortality rates among residents. We used estimates of deaths averted among residents under hypothetical testing scenarios (50%, 75%, 125%, 150%, and 200% of the actual testing volume, as described above) as inputs. We proportionally adjusted the direct and direct- overhead costs of these hypothetical reduced and increased testing volumes. We used the QALY value for a COVID-19 death to calculate the QALYs gained due to deaths averted under the hypothetical testing scenarios compared with actual testing rates. The time horizon of the analysis was consistent with the evaluation period. We also performed a sub-analysis for the two financial years (FY) within the study period: FY21, comprising the six-month period from October 2020 to March 2021; and FY22, comprising the full financial year from April 2021 to March 2022.

We present the incremental cost-effectiveness ratio (ICER) for the cost per death averted in residents due to testing and the cost per QALY gained in residents due to testing. All costs and results were collected and are presented in pound sterling (GBP). More details on our approach, input parameters, and sources can be found in the Supplementary Materials Section 11.

## Results

### Characteristics of care homes studied

Care homes whose primary clients were older people (7,020; 47·4%), people with learning disabilities (4,233; 28·6%), or people with dementia (1,881; 12·7%) accounted for more than 88% of the organisations in our dataset. A substantial proportion of care homes were residential homes (10,458; 70·6%) and were CQC-rated “good” (11,197; >75%). Care homes whose primary clients had learning or physical disabilities had on average, lower numbers of staff per resident compared with care homes serving older people and those with dementia (Table 1).

### Intended and reported PCR and LFD testing intensities

Testing policies for the adult social care sector in England evolved throughout our evaluation period (Supplementary Table S1 and Supplementary Materials Section 1). Whereas the reported testing intensities of PCR tests per staff member and per resident remained consistently at or near the guidance levels, the reported testing intensities of LFDs among staff were consistently lower than the intended testing rates (Fig. 2(A), Supplementary Figures S1 and S2). The median number of reported tests per resident or staff member per week was higher in better CQC-rated care homes, i.e. those rated “outstanding” or “good”, and the level of staff testing was highest in care homes serving older individuals and individuals with dementia.

**Fig. 2.**
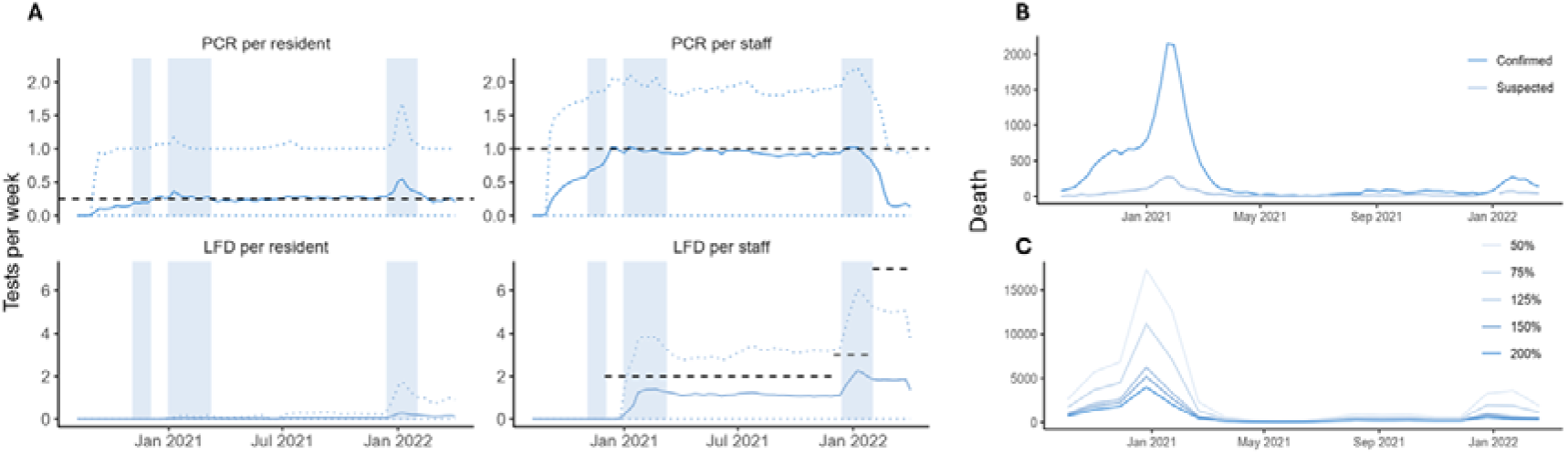
(A) Average reported numbers of tests per person per week in England for adult social care staff and residents. Blue vertical bars show periods when care homes were closed to visitors (with the exception of visiting for palliative care/end-of-life and named essential caregiver visits) due to government closure policies and community-wide lockdowns. (B) Confirmed and suspected COVID- 19-related deaths of residents in 14,805 English care homes studied. (C) The model-projected total COVID-19-related deaths under hypothetical testing levels relative to historical levels. Each line corresponds to a different counterfactual scenario when the numbers of tests were at the levels shown on the right-hand side of the panel relative to the historical levels, e.g. 50% means that the number of tests was decreased by 50% from its factual level.

### Volumes of tests and associated costs

In total, 227 million LFDs were distributed from October 2020 to March 2022 for the adult social care testing service, representing 11·4% of the total number distributed in England (Supplementary Table S2). There were 41 million PCR tests registered for adult social care through Pillar 2, representing 35·5% of total Pillar 2 PCR tests in England. The total financial cost of the adult social care testing service (LFD and Pillar 2 PCR) was estimated to be GBP 4·8 billion, representing 20·6% of England’s total testing spend (Supplementary Table S3). The average unit cost in the adult social care testing service for an LFD and a PCR test was GBP 6·38 and 78·30, respectively.

### Key drivers of changes in reported testing intensity

Only some testing policy changes were associated with changes in reported testing intensities among staff. The introduction in December 2020 of twice-weekly asymptomatic LFD testing among staff in care homes was associated with an average increase in reported LFD testing intensity of 0·10 tests per staff member per week (95% confidence interval (CI), 0·06–0·15). The move to daily LFD testing in February 2022 was also associated with an increase in reported LFD testing intensity of approximately 0·25 tests per staff member per week on average (95% CI, 0·21–0·30). Two other policy changes we considered were not associated with changes in staff testing intensity (Supplementary Table S4).

Care home resident testing tended to increase if positive cases were found. In several care homes, reported testing intensities appeared to change rapidly, coinciding with or following increases in testing positivity (Supplementary Figure S3, Supplementary Table S4-S5, and Supplementary Materials Section 1); this likely indicated a shift to outbreak testing, a policy introduced in February 2021 aimed at controlling outbreaks once they were discovered. (Here, an outbreak was defined as two or more positive (or clinically suspected), linked cases that occurred in the same setting within a 14-day period.) This observation was supported by regression analyses showing that higher numbers of positive results per care home member (including residents and staff members) were associated with increased reported testing intensities in residents the following week (Table 2)

**Table 2.** Regression results for determinants of outbreak size during outbreak discovery and outbreak control and for resident COVID-19-related deaths. The outbreak discovery and control model results correspond to generalised linear models estimated using a Poisson likelihood and a log-link function with an offset term. The resident COVID-19-related mortality model results correspond to generalised linear models using a negative binomial likelihood and a log-link function with an offset term as described in the Methods section. Coefficients and 95% confidence intervals are shown on the exponentiated scale when applicable. Estimates of the weekly time dummies and the COVID-19 prevalence effects are both suppressed for readability.

| Category | Variable | Outbreak discovery |  | Outbreak control |  | Resident mortality |  |
| --- | --- | --- | --- | --- | --- | --- | --- |
|  |  | Resident PCR + LFD positive counts | Staff PCR + LFD positive counts | Resident PCR + LFD positive counts | Staff PCR + LFD positive counts | Total resident COVID-19-related deaths | Confirmed resident COVID-19-related deaths |
| Testing intensity | log (0.1 + test intensity in residents (lag=1)) | 0.926***<br>(0.916, 0.936) | 0.999<br>(0.994, 1.005) | 0.675***<br>(0.669, 0.681) | 0.894***<br>(0.889, 0.898) |  |  |
|  | log (0.1 + test intensity in staff (lag=1)) | 0.745***<br>(0.736, 0.755) | 0.662***<br>(0.655, 0.668) | 0.729***<br>(0.721, 0.738) | 0.488***<br>(0.481, 0.495) |  |  |
| Infections in care home | Number of positive results in residents |  |  |  |  | 0.997**<br>(0.995, 1.000) | 0.998<br>(0.996, 1.001) |
| CQC rating (baseline: Inadequate) | Requires improvement | 0.990<br>(0.925, 1.056) | 0.998<br>(0.952, 1.044) | 0.971<br>(0.930, 1.012) | 0.917***<br>(0.881, 0.953) | 1.061<br>(0.894, 1.228) | 1.170*<br>(0.988, 1.353) |
|  | Good | 0.904***<br>(0.840, 0.967) | 0.948**<br>(0.904, 0.993) | 0.959**<br>(0.919, 0.999) | 0.913***<br>(0.878, 0.948) | 1.043<br>(0.881, 1.204) | 1.155<br>(0.978, 1.332) |
|  | Outstanding | 0.784***<br>(0.707, 0.861) | 0.857***<br>(0.807, 0.907) | 0.907***<br>(0.861, 0.953) | 0.894***<br>(0.856, 0.932) | 1.005<br>(0.815, 1.195) | 1.121<br>(0.914, 1.327) |
|  | No record | 0.876*<br>(0.732, 1.020) | 0.935<br>(0.843, 1.028) | 0.962<br>(0.873, 1.051) | 0.954<br>(0.888, 1.020) | 1.086<br>(0.721, 1.451) | 1.261<br>(0.874, 1.648) |
| Primary clients | Older individuals (≥65 years) | 1.196***<br>(1.159, 1.233) | 0.888***<br>(0.868, 0.909) | 1.318***<br>(1.295, 1.342) | 1.049***<br>(1.033, 1.066) | 2.200*** (2.004, 2.395) | 2.382***<br>(2.168, 2.597) |
|  | Individuals with dementia | 1.251***<br>(1.210, 1.292) | 0.929***<br>(0.905, 0.952) | 1.303***<br>(1.278, 1.328) | 1.056***<br>(1.038, 1.074) | 2.208*** (2.009, 2.408) | 2.336***<br>(2.118, 2.555) |
|  | Individuals with learning disabilities | 1.246***<br>(1.202, 1.290) | 1.199***<br>(1.176, 1.222) | 0.962**<br>(0.931, 0.992) | 0.916***<br>(0.896, 0.936) | 0.254***<br>(-0.003, 0.510) | 0.278***<br>(0.001, 0.555) |
| Local proportion of variant of concern (baseline: Alpha) | Delta | 1.285<br>(0.698, 1.872) | 0.999<br>(0.743, 1.256) | 0.507**<br>(-0.116, 1.131) | 0.455***<br>(-0.017, 0.928) | 4.367<br>(1.726, 7.009) | 5.957<br>(3.198, 8.716) |
|  | Omicron | 1.350***<br>(1.197, 1.504) | 1.319***<br>(1.223, 1.414) | 0.936*<br>(0.864, 1.007) | 1.095***<br>(1.041, 1.149) | 0.871 (0.372, 1.371) | 0.861<br>(0.305, 1.416) |
| Care home size | Care workers per resident (staff-to-resident ratio) | 1.037***<br>(1.012, 1.063) | 1.031***<br>(1.017, 1.045) | 0.699***<br>(0.683, 0.715) | 0.568***<br>(0.556, 0.580) | 0.904*** (0.849, 0.959) | 0.900***<br>(0.841, 0.959) |
|  | Fraction of agency workers | 1.077***<br>(1.042, 1.112) | 1.071***<br>(1.040, 1.101) | 1.010<br>(0.982, 1.038) | 0.959***<br>(0.934, 0.984) | 1.137**<br>(1.038, 1.236) | 1.134**<br>(1.028, 1.240) |
|  | Total resident count | 0.981***<br>(0.980, 0.982) | 0.980***<br>(0.980, 0.981) | 1.003***<br>(1.003, 1.003) | 1.005***<br>(1.004, 1.005) | 0.999<br>(0.996, 1.002) | 0.999<br>(0.996, 1.003) |
| Care home type | Community care home | 0.983<br>(0.930, 1.036) | 0.937***<br>(0.909, 0.966) | 1.022<br>(0.993, 1.052) | 1.026***<br>(1.007, 1.046) | 0.858**<br>(0.725, 0.991) | 0.862**<br>(0.720, 1.003) |
|  | Acute care home | 2.233**<br>(1.491, 2.975) | 1.308<br>(0.858, 1.759) | 1.103<br>(0.559, 1.647) | 0.870<br>(0.420, 1.320) | Note1 | Note1 |
|  | Nursing home | 0.915*** | 0.939*** | 0.935*** | 1.040*** | 1.247*** | 1.235*** |
|  |  | (0.894, 0.936) | (0.926, 0.952) | (0.923, 0.947) | (1.030, 1.049) | (1.202, 1.292) | (1.187, 1.283) |
|  | Independent organisation | 1.036<br>(0.607, 1.465) | 1.070<br>(0.889, 1.251) | 1.089<br>(0.895, 1.282) | 1.003<br>(0.912, 1.094) | 1.847*<br>(1.140, 2.555) | 1.804<br>(1.055, 2.552) |
| Care home vaccination coverage | Average number of all vaccine doses per resident | 0.990***<br>(0.986, 0.994) | 1.001***<br>(1.000, 1.001) | 0.985***<br>(0.982, 0.987) | 0.999*<br>(0.998, 1.000) | 0.991<br>(0.979, 1.003) | 0.994<br>(0.982, 1.006) |
|  | Average number of all vaccine doses per staff | 0.993***<br>(0.990, 0.996) | 0.988***<br>(0.986, 0.990) | 0.994***<br>(0.992, 0.995) | 1.00<br>(0.999, 1.001) |  |  |
| Constant |  | 7.922***<br>(7.777, 8.066) | 13.611***<br>(13.527, 13.695) | 0.391***<br>(0.241, 0.541) | 0.458***<br>(0.337, 0.580) | 0.025***<br>(-0.422, 0.473) | 0.020***<br>(-0.452, 0.492) |

Note:
<sup>1</sup> Estimates of risk of COVID-19-related deaths in acute care homes were dropped from the output as they had extremely high uncertainty reflecting the low numbers of such care homes in our dataset.
<sup>2</sup> \*p<0.1; \*\*p<0.05; \*\*\*p<0.01; p without \* means p>0.1

### Key drivers of outbreak discovery and control in care homes

Higher reported testing intensities among staff and residents were associated with smaller outbreak sizes before outbreaks were detected and while outbreaks were forming, suggesting earlier outbreak discovery and more rapid outbreak control. In our model, the number of positive cases of residents and staff uncovered during outbreak discovery was 0·67% (95% CI, 0·57–0·76) and 2·53% (95% CI, 2·42–2·63) lower, respectively, if the testing intensity (including PCR and LFDs) among staff increases 10% (Table 2). The number of positive cases among staff uncovered during outbreak control was 3·36% (95% CI, 3·28–3·36) lower if the testing intensity (including PCR and LFD tests) among staff increases 10%, while the corresponding figure in residents was higher, at 6·05% (95% CI, 5·34–6·17) (Table 2).

Nursing and care homes with better CQC ratings experienced smaller outbreaks when initially uncovered, that were more rapidly controlled, compared with homes with a lower CQC rating. Outbreaks uncovered among residents in care homes with an “outstanding” CQC rating were, on average, 21·6% (95% CI, 13·9–29·3) smaller than those uncovered in care homes rated “inadequate” by CQC; the corresponding figure for staff outbreaks was 14·3% (95% CI, 9·3–19·3) (Table 2). During the outbreak control phase, care homes rated “outstanding” had an average 9·3% (95% CI, 4·7–13·9) reduction in weekly numbers of positive cases among resident vs “inadequate” care homes; the corresponding figure for positive cases among staff in homes rated “outstanding” was 10·6% (95% CI, 6·8–14·4) (Table 2).

The number of positive cases discovered among residents of nursing care homes during outbreak discovery was 8·5% (95% CI, 6·4–10·6) lower compared with non-nursing care homes, while the corresponding number was 6·1% (95% CI, 4·8–7·4) among staff (Table 2). Once an outbreak occurred, it was brought under control more rapidly among residents in nursing homes compared with care homes supporting other types of residential clients; however, the same was not true among staff (Table 2).

Large care homes tended to have smaller outbreaks when they were initially uncovered but they were more difficult to control once detected. If the total resident count increased by one member, the number of positive cases among residents and staff in discovered outbreaks was associated with a decrease of 1·9% (95% CI, 1·8–2·0) and 2·0% (95% CI, 1·9– 2·1), respectively (Table 2). However, during an outbreak, a one-unit increase in the number of residents was associated with a reduction in the rate of outbreak control by 0·3% and 0·5% among residents and staff, respectively (Table 2).

Variants of concern were associated with initial outbreak size and the ability of care homes to control outbreaks. Positive cases uncovered among residents during outbreak discovery were on average 35·0% (95% CI, 19·7–50·4) higher during the Omicron variant phase compared with the Alpha variant phase, while the corresponding number was 31·9% (95% CI, 22·3–41·4) among staff (Table 2). The Omicron variant phase tended to be associated with a lower ability of care homes to control outbreaks among staff, and positive cases among staff during the Omicron variant phase were on average 9·5% (95% CI, 4·1– 14·9) higher than during the Alpha variant phase (Table 2).

Higher staff-to-resident ratios were associated with larger outbreak sizes when they were uncovered but outbreak control was more rapid thereafter. We estimated that an increase of one additional staff member per resident led to a 3·7% (95% CI, 1·2–6·3) increase in positive cases uncovered among residents and a 3·1% (95% CI, 1·7–4·5) increase among staff (Table 2). However, having more staff per resident was associated with marked reductions in the size of outbreaks subsequent to their discovery in both residents (30·1%; 95% CI, 28·5–31·7) and staff (43·2%; 95% CI, 42·0–44·4).

Outbreaks in care homes serving older individuals or those with dementia were controlled more slowly than in other types of care homes. The numbers of positive cases uncovered during outbreak discovery among residents in care homes whose primary clients were individuals who were older (≥65 years), had dementia, or had learning disabilities were 19·7% (95% CI, 15·9–23·3), 25·1% (95% CI, 21·0–29·2), and 24·6% (95% CI, 20·2–29·0) higher, respectively, compared with care homes that served different clients. Once detected, outbreaks among residents were brought under control more slowly in care homes whose primary clients were older individuals (≥65 years) (31·8% lower; 95% CI, 29·5–34·2) or who had dementia (30·3% lower; 95% CI, 27·8–32·8) than in care homes with other client groups. Positive cases among residents declined more rapidly after outbreaks were uncovered in care homes whose primary clients were individuals with learning disabilities (3·8%; 95% CI, 0·8–6·9) compared with care homes whose clients did not have learning disabilities (Table 2).

Among staff, the number of positive cases uncovered during outbreak discovery at care homes whose primary clients were older individuals (≥65 years) or those with dementia were 11·2% (95% CI, 9·1–15·9) and 7·1% (95% CI, 4·8–9·5) lower, respectively, compared with care homes whose primary clients were not in these groups. Once outbreaks were detected among staff, they took longer to control in care homes whose primary clients were older individuals (≥65 years) or those with dementia (4·9% (95% CI, 3·3–6·6) and 5·6% (95% CI, 3·8–7·4), respectively). However, outbreak control was 8·4% (95% CI, 6·4–10·4) faster among staff in care homes whose primary clients were individuals with learning disabilities (more results of sensitivity analysis for staff and resident testing on outbreak discovery and outbreak control can be found in Supplementary Table S6-S14).

### COVID-19-related death in care homes and its key drivers

There were 24,500 COVID-19-related deaths of residents, from 14,805 care homes in England, reported to CQC during our evaluation period. The distribution of these deaths indicates large heterogeneities depending on the characteristics of the care homes studied (Table 2), suggesting care homes had various risk factors for the likelihood of COVID-19- related deaths. The highest peak in COVID-19-related confirmed and suspected deaths occurred in February 2021, with more than 2000 deaths reported per week (Fig. 2(B)). The primary client of a care home was the strongest determinant of the magnitude of COVID-19- related deaths.

Supplementary Figure S3 shows that, in those care homes primarily serving older patients (≥65 years) or those with dementia, there was a strong positive association between COVID- 19-positive cases and deaths, likely reflecting the underlying (and well-documented) susceptibility of these populations to severe COVID-19 outcomes^50^ ^51^. Care homes primarily serving individuals with learning disabilities or mental health issues had a lower risk of COVID-19-related deaths. These associations were also reflected in our regression results: the risk of COVID-19-related death (including both confirmed and suspected COVID-19- related death) after testing positive was increased by 120·0% (95% CI, 100·0–140·0) in care homes whose primary clients were older individuals (≥65 years) or were individuals with dementia. In comparison, the risks of COVID-19-related death in care homes whose primary clients were individuals were individuals with learning disabilities were 74·6% (95% CI, 49·0– 100·0) lower compared with care homes catering for other types of clients.

Nursing homes had a higher rate of COVID-19-related deaths compared with non-nursing homes, with a risk of COVID-19-related death after testing positive that was 24·7% (95% CI, 20·2–29·2%) higher (Table 2). Higher staff-to-resident ratios were associated with a reduction in the risk of COVID-19-related deaths. We found that the more care workers per resident, the lower the risk of death from a given positive result. The median number of staff per resident in our dataset was 0·93, and our models indicate that a 25% increase in this ratio would be associated with an approximately 2·4% decrease in the risk of death (95% CI, 1·0–3·8; Table 2).

Next, we projected mortality in care homes under counterfactual testing scenarios. Our model indicated that with a reported testing intensity at 50% of the true levels, deaths among residents of care homes in England would increase by 32,160 (uncertainty interval (UI), 27,200–37,740), a 129% increase (UI, 109–152%) in COVID-19-related deaths. With testing at 75%, we estimated an increase of 11,910 (UI, 8500–15,700) and a corresponding percentage change in deaths of 48% (UI, 34–63%). We estimated an increase in reported testing intensity of 25% would have reduced deaths by 4680 (UI, 2270–6810), 18% (UI, 9– 27%) of overall deaths averted (Fig. 2(C); see sensitivity analysis results in Supplementary Figures S4 and S5).

### Testing in care homes can be considered highly cost effective

Table S15 illustrates the cost effectiveness of the actual testing levels using various comparators (see Supplementary Table S16 for the complete set of cost-effectiveness results). We estimated that there was a cost saving of GBP 38,300 per death averted compared with a lower testing rate of 50% of the actual rates. If testing had been conducted at increased levels of 150% and 200% of actual levels, our model indicated that it would have saved GBP 154,100 and 210,000, respectively, per additional death that could have been averted. In terms of QALYs, the model estimates that the actual testing had a cost of GBP 5700 per QALY gained compared with a testing rate of 50% (of the actual rate), corresponding to a highly cost-effective strategy at both HM Treasury’s willingness to pay threshold of GBP 70,000 per QALY gained^52^ and the National Institute for Health and Care and Excellence (NICE) threshold of GBP 30,000.^53^ Furthermore, the model predicted that if testing had been increased to 150% and 200% of actual levels, it would have cost GBP 22,700 and 31,000, respectively, for each additional QALY that could have been gained. This suggests that increasing testing of residents and staff to up to double the actual volume may also have been a cost-effective intervention, although the degree of cost effectiveness would have diminished.

While cost effective throughout, testing in care homes was substantially more cost effective prior to the full vaccination rollout. On average, we estimated that testing in care homes was 3·5-times more cost effective prior to vaccination rollout for the scenarios considered. Fig. 3 illustrates the costs per QALY gained at various testing rates in care homes by financial year. Testing in care homes was cost effective compared with lower levels of testing at 50% and 75% of the actual volume for both FY21 (half year) and FY22 (full year). If testing had been increased to levels of 125%, 150%, or 200% of the actual testing, the additional QALYs that could have been gained would also have been cost effective for FY21 and may have been cost effective for FY22 at 125% or 150%. The incremental cost per QALY gained for FY21 was substantially lower than for FY22, indicating that testing in care homes was more cost effective prior to vaccination rollout. FY22 aligned with the rollout of vaccination; by April 2022, the start of FY22, 85% of those aged ≥65 years had received their first dose of the vaccine.

**Fig. 3.**
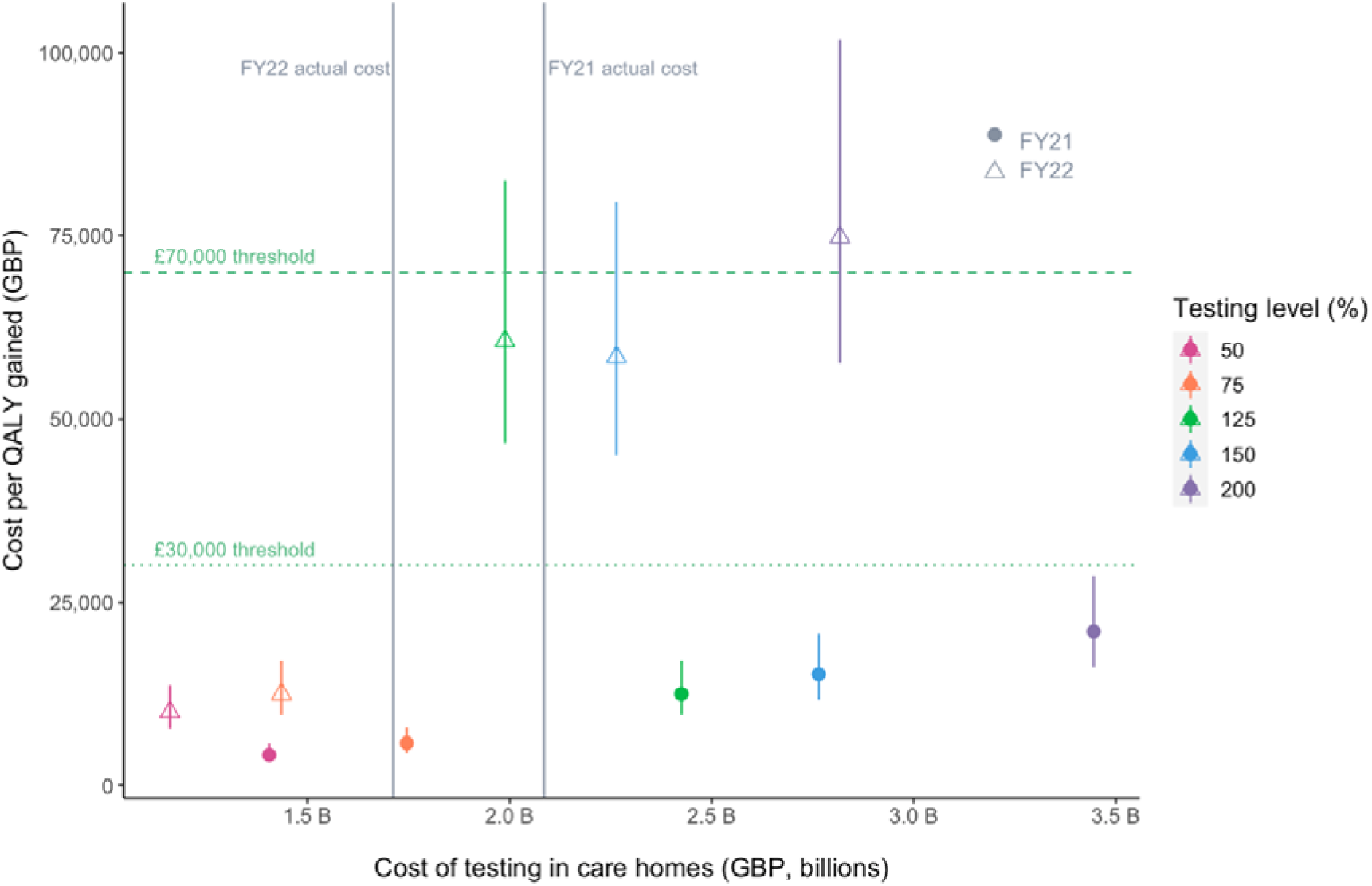
Cost effectiveness of actual testing levels in adult social care homes in England compared with various hypothetical levels of testing. The whiskers represent the cost-effectiveness ranges using a minimum QALY of 4.98 per death averted and a maximum of 8.8 per death averted. The points falling below the GBP 70,000 willingness to pay threshold are considered to be cost effective. The points to the right of the vertical lines indicate testing volumes that would be more expensive than the actual strategy was (125%, 150%, and 200% of the actual testing volume).

## Discussion

Our study, the first large-scale observational evaluation of the impact and cost effectiveness of PCR and LFD testing for COVID-19 in care homes in England, indicated that testing of residents and staff effectively contained COVID-19 outbreaks and was cost effective in saving the lives of residents. For future pandemics with similar at-risk populations, it will be essential to rapidly develop reliable diagnostic tests and deploy them promptly to care homes, while shielding particularly vulnerable individuals until these tests become available.

Our evaluation of testing implementation found that reported testing intensities responded partially to changes in policy over time, with considerable heterogeneity among care homes.

Testing intensities for LFDs remained consistently below intended levels among staff from October 2020 to March 2022. Contributing to this implementation shortfall, in addition to poor adherence to testing policy among staff,^54^ included issues of test supply during the early stages of the pandemic^55^ and under-reporting of implemented tests, especially LFDs, which reporting reliant on self-reporting (whereas PCR tests were conducted in and reported by a laboratory). Individuals may have perceived a negative test result to be unimportant and not worth reporting, while positive tests may have been withheld out of fear of losing work income^19^. Higher testing intensities were associated with earlier outbreak discovery and more rapid outbreak control. A similar finding was reported for US nursing homes, with the number of COVID-19 cases per 100 potential outbreaks 71·5% lower in high-testing facilities than in low-testing facilities.^56^

An innovation in our study was the decomposition of the process of outbreak mitigation into outbreak discovery and outbreak control; we found certain characteristics of care homes may play different roles in these two processes. Our findings are largely consistent with the literature that does not separate these activities but assesses the correlates of outbreaks in care homes overall^12^ ^33^ ^57^. We found that large care homes tended to have smaller outbreaks when they were initially uncovered but that these outbreaks became more difficult to control subsequent to their detection. Likewise, in our study higher staff-to-resident ratios were associated with larger outbreak sizes when these were uncovered but with more rapid outbreak control afterwards, consistent with both a previous UK report^12^ and a study from Ireland.^29^

In contrast with our findings, an audit found that during the first wave of the COVID-19 pandemic (March–June 2020), CQC ratings (different from those we used) were not associated with COVID-19 outbreaks or asymptomatic cases in Liverpool care homes.^31^ However, a similar link between the severity of care home outbreaks and care quality standards has been reported for care homes in the US.^58^ ^59^

Our results are aligned with those of a population analysis of 189 care homes in the NHS Lothian health region, in Scotland, which estimated that the size of care homes for older people was strongly associated with an increased likelihood of a COVID-19 outbreak (odds ratio per 20-bed increase, 3·35; 95% CI, 1·99–5·63).^6^ We found that outbreaks in homes serving older individuals (≥65 years) or those with dementia were controlled more slowly.

We identified that testing in care homes was highly cost effective throughout our evaluation period and 3·5-times more cost effective prior to vaccination rollout. Although the rollout of vaccination may have made ongoing testing appear marginally less cost effective, continued testing should be considered a reasonable and overall cost-effective insurance strategy given the uncertainties around the long-term effectiveness of vaccines and the potential for future variants of concern. This also raises the question of whether a standard health economics approach is appropriate for analyses such as this, post-vaccination rollout. In England, testing was continued based on the precautionary principle and because the potential impact of new variants was unknown. Traditional cost-effectiveness analyses do not take into account the willingness to pay for this type of assurance.

There are strengths and weaknesses to our study. We had access to large-scale testing data covering all PCR and LFD tests within 14,805 care homes in England reported through the Pillar 2 testing programme between October 2020 and March 2022. Testing data were linked to care home characteristics, test costs, and resident mortality information to contextualise drivers of testing patterns and resident mortality. However, severe COVID-19 infection can substantially reduce an individual’s quality of life even if it does not lead to death, but due to a lack of hospitalisation data our economic analysis did not include potential cost savings from averted hospitalisations. If these were considered, testing in care homes would likely have emerged as even more cost effective. We found that testing was effective at containing outbreaks in staff, but without staff hospitalisation and deaths data, we could not investigate whether testing could prevent these severe infection outcomes, even though staff in similar settings have been shown to be at high risk of exposure and severe infection^60^ ^61^.

Our study had some data-based limitations. Whilst our data records deaths in care homes, we do not have information on the location of patients when they died, and it is possible that some of these deaths could have been due to hospital-acquired infections. The dynamic occupancy of care home workers and presence of residents were not comprehensively available, so interpolation analyses were used to impute the missing data (see more details in Supplementary Materials Section 12). These highlight the challenges of accessing relevant datasets for care homes and underscores the importance of ensuring such data are recorded and can be made accessible. Additionally, there are wider benefits and costs of testing which could not be probed in our study because they are challenging to quantify.

These include issues such as the effects of testing on end-of-life quality, the costs of isolation and the resulting mental distress experienced by individuals in care homes due to reduced visitations. In addition, averting hospitalisations will have freed up hospital and staff capacity, enabling better care for emergencies and other diseases.

To produce the mortality projections, we assumed that our regression models captured the causal impacts of testing policy on outbreak discovery and control and also captured the link between positive test counts and deaths. Our models included a large range of hypothesised confounders, however, determining causality from these observational data is inherently challenging. More linked data, connecting patient test results with health outcomes could further elucidate these connections.

The impact of COVID-19 testing in various settings, e.g. nursing homes^33–36^, schools^62^ ^63^, healthcare facilities^64^ ^65^ and the general population^66^ ^67^, has been extensively explored in the mathematical modelling literature. For example, simulations of an agent-based model indicated that screening testing in nursing homes could reduce COVID-19 mortality in the U.S. when COVID-19 community incidence was high and/or booster uptake was low^36^ ^68^.

Another study reported that when combined with high-quality infection control practices, outbreak testing could be an effective approach to preventing COVID-19 in nursing homes, particularly if optimized through increased test frequency and use of tests with rapid turnaround^34^ ^69^.

High attack rates and repeated infections of COVID-19 in care homes in England were reported even after COVID-19 vaccination^69^ ^70^. Immunological studies showed strong peak immunogenicity but rapid antibody waning following a third vaccine dose in older residents of care homes^71^, highlighting the continued vulnerability of care homes serving such individuals. COVID-19 is not the only infectious disease threatening residents’ lives. Care homes have long suffered from various life-threatening but often neglected respiratory diseases pre-pandemic^72^ ^73^, and recent studies show that more crowded nursing homes have higher incidences of respiratory infection and mortality^74^. Educational interventions to improve infection control procedures and compliance by staff and other antiviral prophylaxis therapies have been recommended to reduce the transmission of respiratory infections in care homes^75^.

This study has implications for the development of testing policies in care homes for future pandemics, particularly for those infectious diseases which have higher morbidity and mortality in older adults or adults with similar characteristics to care home residents. Our analysis found that while testing was a crucial public health intervention in care homes, there were also large heterogeneities across care homes. This raises questions about whether a one-size-fits-all policy for care home testing would be most effective in the event of a future pandemic. It is clear, however, that the impact we have outlined here could not have been achieved through the outbreak management alone that was in place to tackle disease outbreaks in care homes prior to the pandemic, and the testing programme was essential in achieving this impact. Our results indicate testing remained cost effective even when there were large increases in testing, and in a future pandemic, targeting even higher testing intensities for the most at-risk care homes should be considered. Post-vaccination rollout, the precautionary principle suggests that testing be continued; however, efforts should be made to evaluate the willingness to pay for this type of assurance.

## Data Availability

The data supporting the findings of this study are available within the paper and its supplementary materials. The data were made available by UKHSA to the manuscript's authors as part of a retrospective evaluation of England's COVID-19 testing programme. The authors cannot make the underlying datasets publicly available for ethical and legal reasons, particularly given the sensitive nature of the information included. Applications for access to the anonymised data should be submitted to UKHSA.

## Contributorship statement

SC, RC, RH, LW, RS, BL, TF, and SH conceived and designed the study. LW, MV, RS, RN and BL supervised the work. SC, RC, RH, LC, SB, PD, LHT, OK and RN collected the testing data. SC and MA collected the testing policy data. RH and RS collected the costing data. SC, RC, RH, LC, RS and BL coded the models and analysed the data. SC, RC, MV, LW, KS and BL advised on the methodologies. SC, RC, RH, and BL wrote the initial draft of the manuscript. All authors in EY-Oxford Health Analytics Consortium listed below contributed to guiding the research questions, interpretation of results, and reviewing and approving the manuscript. The data were restricted due to privacy concerns and only available to a subset of the authors. SC and BL accept full responsibility for the finished work and the conduct of the study. BL had access to the data and controlled the decision to publish.

EY-Oxford Health Analytics Consortium membership list:

Ricardo Aguas, Ma’ayan Amswych, Billie Andersen-Waine, Sumali Bajaj, Kweku Bimpong, Adam Bodley, Liberty Cantrell, Siyu Chen, Richard Creswell, Prabin Dahal, Sophie Dickinson, Sabine Dittrich, Tracy Evans, Angus Ferguson-Lewis, Caroline Franco, Bo Gao, Rachel Hounsell, Muhammad Kasim, Claire Keene, Ben Lambert, Umar Mahmood, Melinda Mills, Ainura Moldokmatova, Sassy Molyneux, Reshania Naidoo, Randolph Ngwafor Anye, Jared Norman, Wirichada Pan-Ngum, Sarah Pinto-Duschinsky, Sunil Pokharel, Anastasiia Polner, Katarzyna Przybylska, Emily Rowe, Sompob Saralamba, Rima Shretta, Sheetal Silal, Kasia Stepniewska, Joseph Tsui, Merryn Voysey, Marta Wanat, Lisa White, Gulsen Yenidogan

## Data sharing statement

The data we used in this study on tests, vaccination, care home deaths, NECS Capacity Tracker, and testing costs were made available by UKHSA and CQC to the manuscript’s authors as part of a retrospective evaluation of England’s COVID-19 testing programme. Although these datasets cannot be released publicly due to the sensitive nature of the information included, applications for access to the anonymised data should be submitted to UKHSA. Weekly estimates of community prevalence were obtained using a debiasing methodology, with results published in reference^2^.

## Ethics statement

The study protocol for the evaluation project, which this research fed into, was granted ethics approval by the UKHSA Research Ethics and Governance Group, reference number NR0347. All relevant ethics guidelines were followed throughout.

## Acknowledgements

Medical editing support in the preparation of this paper was provided by Adam Bodley, according to Good Publication Practice. The authors would like to express their gratitude to Oliver Munn, Sarah Tunkel, Nick Sharp, Sariyu Shoge, and Olutoye Olatunbosun of UKHSA for sponsoring this research and enabling access to the data used in this study. SH is supported by the National Institute for Health Research (NIHR) Health Protection Research Unit in Healthcare Associated Infections and Antimicrobial Resistance (NIHR200915), a partnership between the UK Health Security Agency (UKHSA) and the University of Oxford. The views expressed are those of the author and not necessarily those of the NIHR, UKHSA or the Department of Health and Social Care.

